# Efficacy of Intrawound Vancomycin in Prevention of Periprosthetic Joint Infection After Primary Total Knee Arthroplasty

**DOI:** 10.1101/2023.05.05.23289368

**Authors:** Praharsha Mulpur, Tarun Jayakumar, Ramakanth R Yakkanti, Aditya Apte, Kushal Hippalgaonkar, Adarsh Annapareddy, A B Suhas Masilamani, A V Gurava Reddy

## Abstract

**Introduction:** Peri-prosthetic Joint Infection (PJI) after total knee arthroplasty (TKA) is a devastating complication. Intra-wound vancomycin powder has been shown to reduce infection rates in spine surgery. Previous studies on the efficacy of local vancomycin powder in hip or knee arthroplasty are mostly retrospective case series. The aim of this prospective RCT was to evaluate the efficacy and safety of intrawound vancomycin in preventing PJI after primary TKA.

**Methods:** This study was a National Trial Registry-approved RCT of patients undergoing primary TKA. 1022 patients were randomized to the study group (n=507, received 2g intrawound vancomycin powder before arthrotomy closure) and control groups (n=515, no local vancomycin). The minimum follow-up was 12-months. The primary outcome was PJI rate. Secondary outcomes included surgical site infection (SSI) rates, incidence of revision for PJI/SSI, and incidence of wound complications. High-risk groups (Obesity and Diabetes) in both cohorts were also evaluated.

**Results:** The overall infection rate in 1022 patients was 0.66%. There was no significant difference in PJI rate in the study group (0.2%) versus the control group (0.58%), p=0.264. Reoperation rates in the study group (N=4;0.78%) and Control (N=5;0.97%) and SSI rates in the study (N=1;0.2%) and control groups (N=2;0.38%) were comparable. The Vancomycin cohort however demonstrated a significantly higher number of minor wound complications (n=67;13.9%) compared to the control group (n=39;8.4%, p<0.05). There was no difference in PJI/SSI rates or minor surgical complications among high-risk groups and no cases of nephrotoxicity were reported in the study.

**Conclusion:** Intra-wound vancomycin powder does not appear to reduce PJI/SSI rate in primary total knee arthroplasties, including high-risk groups. Although safe from a renal perspective, intra-wound vancomycin was associated with an increase in postoperative aseptic wound complications such as persistent wound drainage. Intra-wound vancomycin may not be effective in reducing the rate of PJI in primary TKA.

## INTRODUCTION

There is an ever-increasing global burden of primary total knee and total hip arthroplasty procedures, with a significant increase in surgery numbers projected by the year 2030 (1–3). With increasing numbers of primary total joint replacement surgeries, healthcare systems across the world are poised to handle an increased burden of revision arthroplasty cases. Worldwide, the reported incidence of PJI after total joint replacement is around 0.8-1.5%, with a 1-2% PJI risk after TKA (4–6).

Periprosthetic joint infection (PJI) is one of the most common indications for revision after TKA (7–9) posing a significant burden to the healthcare system (10,11). There has been increasing interest in the use of local antibiotic powder in the joint before closure to prevent local contamination and biofilm formation. The use of intrawound vancomycin in preventing surgical site infections is well documented in spine surgery (12–14). There are some published reports of the benefits of local vancomycin powder used in surgeries of the elbow, foot and ankle surgery (15,16). However, its use and clinical benefit in total knee arthroplasty is debatable.

The main postulated advantages of using local vancomycin powder are its relatively cheap cost, favourable bacterial spectrum (MRSA, coagulase-negative staphylococci), high local concentrations without systemic adverse effects (17,18). Systematic reviews and meta-analyses on studies evaluating the use of vancomycin in primary total knee or hip arthroplasty reported low-quality evidence with a high risk of study bias (19,20). No randomized control trial (RCT) has evaluated the efficacy of local vancomycin powder in primary TKA.

The primary objective of this study was to evaluate the efficacy of intra-wound vancomycin in reducing SSI or PJI rates in patients undergoing primary total knee arthroplasty. Secondary objectives include the evaluation of wound-healing-related complications, incidence of nephrotoxicity, and the influence of diabetes and obesity on outcomes.

## Methods

This is an Institutional Review Board (IRB) approved prospective randomized, controlled trial (RCT), and was prospectively registered with the National Central Trial Registry (CTRI/2021/02/031310). This is a single-centre RCT of patients from a high-volume tertiary care institute, who underwent primary total knee arthroplasty (TKA) for primary osteoarthritis of the knee, between January 2021 and February 2022. The RCT was a parallel-arm trial with a 1:1 allocation of patients into study and control groups. Study group patients receive intra-articular 2g Vancomycin antibiotic powder before arthrotomy closure, control group patients did not receive local vancomycin powder. The trial was conducted following the guidelines of the Declaration of Helsinki on scientific studies involving human subjects.

All adult patients with primary osteoarthritis of the knee, consenting to primary manual jig-based TKA were eligible for recruitment in this trial. This includes patients with primary OA, with either varus or valgus deformity of the knee. Patients were excluded from the trial if they met any of the following exclusion criteria-refusal to participate in the trial, intra-operative findings suggestive of inflammatory arthropathy or non-specific synovitis, previous knee surgeries, history of intra-articular knee injections within 3 years before surgery, known allergy to Vancomycin, and if they were known cases of chronic immunosuppression (secondary to human immuno-deficiency virus-HIV, malignancy or post-solid organ transplant). Patients who used antibiotics for any cause, within 1-month leading up to surgery were also excluded from the trial.

There were a total of 1208 eligible patients who underwent primary TKA during the study period (January 2021 to January 2022). 56 patients declined to participate in the trial and a further 24 patients were excluded due to exclusion criteria (12 patients used antibiotics for Urinary tract infections, 8 patients with recent intra-articular knee visco-supplementation injections and 4 patients with chronic immunosuppression). After exclusions, 1128 patients were randomized to the study and control groups. 34 patients were excluded from the final data analysis after randomization (16 deaths in the follow-up period, 18 cases with intra-operative findings suggestive of inflammatory arthropathy or non-specific synovitis).

The final study cohorts consisted of 549 patients in the study group and 545 patients in the Control group. There was a loss to follow-up of 42 patients (8%) in the study group and 30 patients (5.76%) in the control group, Final analysis included 507 subjects in the study group and 515 subjects in the control group.

Eligible trial participants undergoing primary total knee arthroplasty were randomized (1:1 allocation by computer randomization) to receive either normal saline lavage with (study group) or without (control group) intra-articular 2g Vancomycin powder prior to arthrotomy closure. All patients were operated on at the same institute, in laminar flow operating rooms. Except for the use of vancomycin powder in the surgical wound, the operating procedures and peri-operative protocols were common to both groups. The patients are blinded and unaware of the assigned groups and follow-up evaluation for infection related complications was performed by a blinded observer.

All cases in this trial received a standard antibiotic prophylaxis protocol, which includes 3 peri-operative intra-venous doses of 1.5g of Cefuroxime. The first dose was administered 1 hour before the skin incision and 2 doses were administered 8 hours and 24 hours after surgery. None of the cases in this trial received prolonged Oral antibiotic use after surgery. Skin preparation at the time of surgery was done with Chlorhexidine skin scrub (3M, Avagard 4% Chlorhexidine Scrub). TKA was performed through a medial para-patellar approach in all cases, under a tourniquet. Tourniquet was inflated from the time of the incision till the cement was fully-set after final implantation. The knee joint was irrigated with a total of 3 L of normal saline with pulse lavage before and after cementing of the implant. All cases received bone cement without antibiotic impregnation (Palacos/ Stryker Simplex low-viscosity cement). Patella was not resurfaced in any case, in either group. After deflation of the tourniquet and haemostasis of bleeding vessels, the study group received 2g vancomycin powder, which was placed into the medial and lateral joint gutters and arthrotomy closed with No.2 Vicryl in an interrupted fashion followed by a continuous STRATAFIX (Barbed-PDS) running stitch. Suction drains were not used in any case.

Post-operative blood investigations (Renal parameters, CBP) were obtained 24 hours after the surgery. In all cases, wound dressing was changed 24 hours later in the PACU, with MEPILEX surgical dressing (*Mepilex Border Post-op, Mölnlycke, Göteborg, Sweden*). Patients in both groups received uniform rehabilitation with assisted walking and range of motion exercises on the first postoperative day. Deep vein thrombosis prophylaxis was common with low-Molecular weight heparin (LMWH) in the immediate postoperative period and Oral Apixaban 2.5mg twice daily for 14 days following surgery. Surgical staple removal is done between Day 14 and Day 21 after surgery. Staple removal after 21 days is considered to be “Delayed staple/clip removal”. Patients were routinely followed up at 2 weeks, 4 weeks, 3 months and 6 months and 12 months after surgery.

Based on the criteria identified by Parvizi et al (21), early peri-prosthetic joint infection was defined as an infection diagnosed within 90 days of surgery. Infection was diagnosed based on the MSIS criteria and the 2018 Definition of Periprosthetic Hip and Knee Infection consisting of 2 positive cultures (from an aspirate and/or at the time of debridement for PJI or SSI), elevated serum ESR, CRP and elevated white cell counts in synovial fluid aspirates (>10,000 cells/mm^3^), PMN percentage greater than 90% in the synovial aspirate(21,22). Cases of possible infections based on the 2018 ICM (International Consensus Meeting) scoring system were confirmed based on the findings of pus in aspirate and intra-operative findings of purulence.

Surgical site infections and deep peri-prosthetic joint infections were considered major surgical complications, necessitating re-operation. Minor complications included delayed wound healing with or without dehiscence, delayed surgical staple removal, and stitch/suture abscess (based on CDC Guidelines) necessitating oral antibiotic use. Persistent wound drainage was defined as, wound drainage beyond 72-hours necessitating surgical dressing change, based on previous definitions of persistent wound drainage (21,23).

Patients were also monitored in the peri-operative period for medical complications of Myocardial infarction, DVT/PE, CVA and nephrotoxicity secondary to the use of vancomycin.

## Statistical analysis

Xu et al conducted a systematic review and meta-analysis of intra-wound vancomycin powder used in primary total knee and hip arthroplasty(24). This meta-analysis concluded that a minimum sample of 1000 patients was required for analysis, to conform to a decrease of PJI rate from 2.74% to 1% in control versus study groups, with a power of 80% and 5% significance level. Assuming a drop-out rate of 10%, we recruited over 1100 eligible subjects for trial participation and randomization. Statistical analysis was performed using a 2-tailed or independent samples t-test for continuous data parameters. The Chi-squared test or Fisher’s Exact test was used for categorical data parameters. Statistical analysis was performed using SPSS Version 24 (International Business Machines-IBM, Armonk, NY). Assuming a power of 80% and 95% confidence intervals, a p-value less than 0.05 was considered significant.

## Results

A total of 1022 patients were included in the final statistical analysis, with 507 patients in the study group and 515 patients in the control group, after exclusions, loss to follow-up and deaths. Demographic variables and baseline characteristics such as comorbidities, BMI, ASA grading, and CCI grading were comparable between both groups and summarised in **Table.1**. The mean tourniquet times and mean haemoglobin drop after surgery were also comparable between both groups. The CONSORT Flowchart of participant recruitment is shown in ***Figure.1***

**Table 1:** Demographic and Baseline Characteristics of the study population

| Characteristics | Study Group<br>N=507 | Control Group<br>N=515 | p-value |
| --- | --- | --- | --- |
| Mean Age (SD) | 61.7 (7.52) | 61.4 (7.39) | 0.520 * |
| Mean BMI (SD) | 28.5 (4.49) | 28.4 (4.52) | 0.722 * |
| Mean Pre-op Haemoglobin (SD) | 12.2 (1.41) | 12.3 (1.33) | 0.243 * |
| Mean Post-op Haemoglobin (SD) | 10.3 (1.34) | 10.4 (1.24) | 0.215 * |
| Mean drop in Haemoglobin (SD) | -1.96 (1.143) | -1.91 (1.111) | 0.478 * |
| Mean ASA (SD) | 2.02 (0.143) | 2.01 (0.186) | 0.336 * |
| Mean Tourniquet Time (min) (SD) | 70.2 (16.35) | 71.1 (21.91) | 0.457 * |
| Smoking N (%) | 58 (11.44) | 60 (11.65) | 0.916 * |
| **CCI Risk Classification** |  |  |  |
| Mild risk | 324 (63.9) | 330 (64.07) | 0.278 † |
| Moderate risk | 156 (30.76) | 164 (31.84) |  |
| High Risk | 27 (5.34) | 21 (4.09) |  |
| Mean CCI (SD) | 2.3 (1.15) | 2.2 (1.14) | 0.163 * |
| Gender Female N (%) | 360 (71) | 358 (69.51) | 0.586 † |
| Male N (%) | 147 (29) | 157 (30.49) |  |
| Diabetics N (%) | 170 (33.53) | 161 (31.26) | 0.438 † |
| Hypothyroidism N (%) | 85 (16.76) | 92 (17.86) | 0.642 † |
| CKD N (%) | 2 (0.39) | 4 (0.77) | 0.424 † |
| ASA N (%) ASA I | 6 (1.2) | 10 (1.94) | 0.220 † |
| N (%) ASA II | 491 (96.84) | 492 (95.53) |  |
| N (%) ASA III | 10 (1.96) | 13 (2.53) |  |

\* independent samples t-test, † Chi-square test

**Fig. 1.**
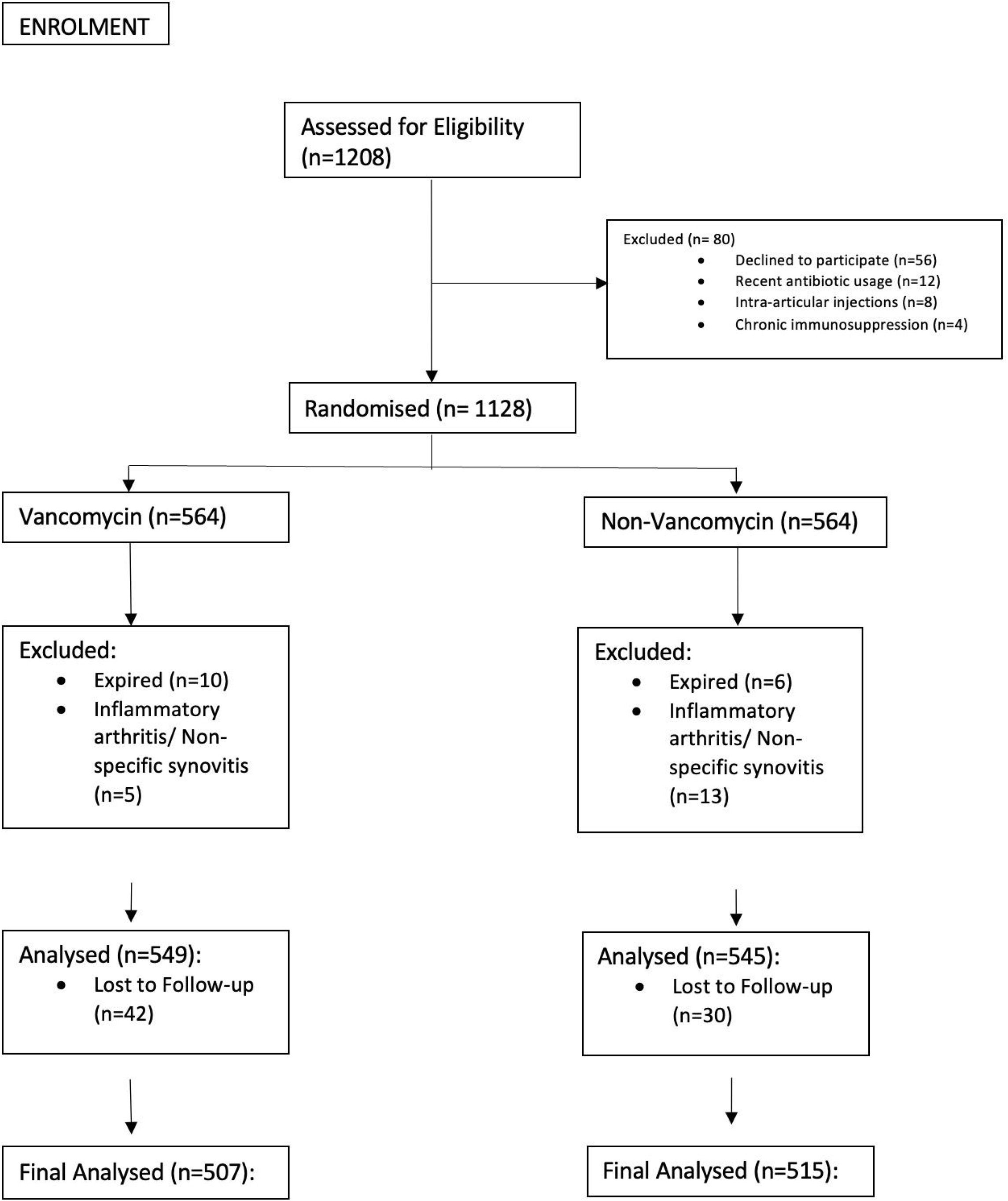

### Infection rates

The overall infection rate in the study population was 0.66%. Periprosthetic joint infection (PJI) was seen in 1 patient (0.19%) in the study group and 3 patients (0.58%) in the control group, and the difference was not statistically significant. Surgical site infection (SSI) was reported in 1 (0.19%) patient in the study group and 2 patients (0.38%) in the control group. 2 patients in the study group had periprosthetic fractures of the distal femur during the follow-up period and underwent ORIF. All complications in the trial are summarized in **Table 2**.

**Table 2:** Summary of complications in the study population

|  | Study Group<br>N(%) | Control Group<br>N(%) | p-value |
| --- | --- | --- | --- |
| **Major Complications** |  |  |  |
| SSI N (%) | 1 | 2 | 0.264 † |
| PJI (DAIR) | 1 | 3 |  |
| Periprosthetic fracture | 2 | 0 |  |
| **Minor Complications** |  |  |  |
| Persistent Wound Drainage | 43 (8.48) | 26 (5.04) | **0.010** † |
| Stitch Abscess | 20 (3.94) | 13 (2.52) |  |
| Delayed stitch removal (>3 weeks) | 36 (7.1) | 36 (6.99) | 0.945 † |
| **Medical Complications** |  |  |  |
| Septic shock and MODS | 0 | 1 | NS † |
| DVT | 0 | 1 |  |
| CVA | 0 | 0 |  |
| Acute kidney injury/<br>Nephrotoxicity | 0 | 0 |  |
| Cardiac complications | 0 | 0 |  |

†Chi-square test

### Peri-prosthetic Joint Infections (PJI)

In the study group, one patient (0.19%) underwent DAIR for a culture-negative PJI, 4 weeks after surgery. This patient had elevated serum ESR, CRP and White cell counts and frank pus aspirated from the joint before DAIR. In the control group, 3 patients (0.58%) underwent DAIR, of which one patient had Staphylococcus aureus isolated, and later went on to have a 2-stage revision due to the persistence of infection. The other two patients had culture-negative PJI and underwent DAIR.

### Surgical Site Infections (SSI)

One patient in the study group presented with an early SSI, with Staphylococcus epidermidis the pathogenic organism isolated. This patient presented with persistent wound discharge which was managed with superficial wound debridement and secondary closure and iv antibiotics. 2 patients in the control group developed SSI, for which one patient underwent debridement and secondary closure for local wound necrosis and the other patient developed septic shock and Multi-Organ Dysfunction (MODS) following wound debridement at an outside hospital, requiring hospitalization and intravenous antibiotics. The major complications of PJI and SSI are summarized in **Table 3**.

**Table 3:**
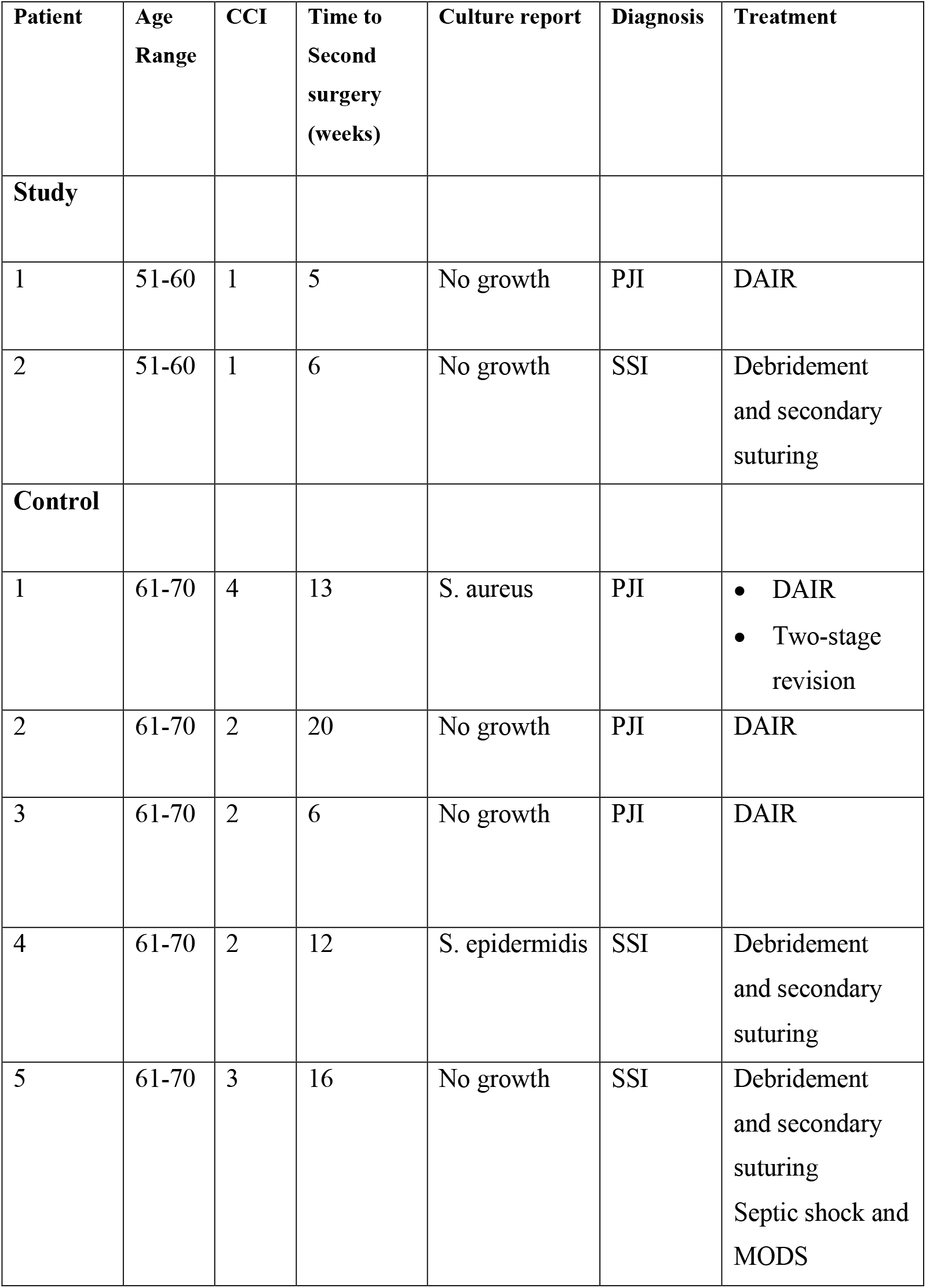
Clinical summary of PJI and SSI cases

### Re-operation Rates

Re-operation rates between both groups were found to be statistically comparable, with 4 0.78%) patients in the study group (1-PJI, 1-SSI, 2-Periprosthetic fracture) versus 5 cases (0.97%) in the control group (3-PJI, 2-SSI). **(Table 2)**

### Wound Complications

The use of vancomycin was associated with a significantly higher number of minor wound complications in the study group (n=67; 13.9%) compared to the control group (n=39; 8.4%, p<0.05). Aseptic wound complications such as wound soakage, maceration and stitch abscess were managed with regular dressings without any additional oral or intravenous antibiotic use. Delayed suture removal (>3 weeks) was found to be similar in both groups and not statistically significant. **(Table 2)**

### Sub-group Analysis

We evaluated differences in complication rates amongst patients of both groups based on BMI and compared diabetics with non-diabetics. There was no difference in PJI/SSI rates or minor surgical complications among high-risk groups, Diabetics versus non-diabetics, **(Table 4)** and high BMI vs normal BMI **(Tables 5 & 6)**.

**Table 4:** Sub-group analysis of outcomes in the high-risk diabetic patients in both groups

|  | Study Group<br>n (%) | Control Group<br>n (%) | p-value |
| --- | --- | --- | --- |
| Number of Diabetics | 170 (33.5) | 161 (31.26) | 0.444 † |
| Mean HbA1C (SD) | 6.6 (2.04) | 6.8 (1.95) | 0.109 * |
| DM-CCI grading |  |  |  |
| Mild | 61 (35.9) | 61 (37.9) | 0.439 † |
| Moderate | 88 (51.8) | 87 (54.0) |  |
| Severe | 21 (12.4) | 13 (8.1) |  |
| Mean CCI (SD) | 3.0 (1.03) | 2.9 (1.14) | 0.141 * |
| Infection rates |  |  |  |
| Major Complications |  |  |  |
| SSI | 0 | 0 | 0.160 † |
| PJI (DAIR) | 0 | 2 |  |
| Minor Complications |  |  |  |
| Maceration, n (%) | 9 (5.3) | 6 (3.5) | 0.329 † |
| Stitch Abscess, n (%) | 15 (9.3) | 4 (2.5) |  |
| Delayed staple removal<br>(>3 weeks) n (%) | 36 (7.5) | 36 (7.3) | 0.942 † |

\* independent samples t-test, † Chi-square test

**Table 5:** Sub-group analysis of minor complications based on the classification of obesity

|  | Study Group<br>Minor complications<br>n (%) | Control Group<br>Minor complications<br>n (%) | p-value |
| --- | --- | --- | --- |
| **BMI grading** |  |  | 0.759 † |
| Normal | 16 (26.2) | 11 (29.7) |  |
| Overweight | 24 (39.3) | 16 (43.3) |  |
| Class I Obese | 15 (24.6) | 5 (13.5) |  |
| Class II Obese | 5 (8.2) | 4 (10.8) |  |
| Class III Obese | 1 (1.6) | 1 (2.7) |  |
| **Total** | 61 (100.0) | 37 (100.0) |  |

† Chi-square test

**Table 6:** Sub-group analysis of major complications based on the classification of obesity

|  | Study Group<br>Major complications<br>n (%) | Control Group<br>Major complications<br>n (%) | p-value |
| --- | --- | --- | --- |
| **BMI grading** |  |  | 0.507 † |
| Normal | 1 (25.0) | 0 (0.0) |  |
| Overweight | 2 (50.0) | 3 (60.0) |  |
| Class I Obese | 1 (25.0) | 2 (40.0) |  |
| Class II Obese | 0 (0.0) | 0 |  |
| Class III Obese | 0 (0.0) | 0 |  |

† Chi-square test

### Systemic Complications and Mortality

The use of intra-wound vancomycin was not associated with acute kidney injury (defined as an increase in serum creatinine levels by more than 0.3mg/dl within 48 hours)(25). A total of 16 patients expired in the follow up period (10 patients in the study group versus 6 patients in the control group. One patient in the control group developed DVT with Pulmonary embolism 45 days post-surgery and expired. Other patients expired secondary to non-surgical factors (1 patient secondary to complications of CKD in the control group, 1 patient with Pleural effusion, 4 patients expired after treatment for trauma unrelated to the prior knee arthroplasty). No cases of CVA, AMI, Nephrotoxicity, Ototoxicity or anaphylactic reactions were observed or documented in the post-operative 90-day follow-up period.

## Discussion

This randomised control trial did not demonstrate any additional clinical benefit of topical or intra-wound vancomycin powder in preventing SSI/PJI in patients undergoing primary total knee arthroplasty. On the contrary, there was a significant increase in wound healing complications in the study group, with a significantly higher incidence of persistent wound discharge and stitch abscess compared to the control group.

PJI after TKA bears a heavy toll on the patient and contributes to the economic burden on the healthcare systems (6,11). The use of local antibiotic powder in orthopaedic surgery is not new, with several reports of benefits in spine surgery (12–14,19,26). However, most of these studies were retrospective and non-randomised. Tubaki et al published the only RCT on the use of vancomycin in spine surgery and concluded that there was no difference in infection rates with the use of vancomycin (27).

The role of topical or intra-wound vancomycin in primary total knee arthroplasty is undecided, with studies both supporting and refuting the efficacy of vancomycin in reducing post-operative PJI rates.

Heckman et al (28) published a meta-analysis and systematic review of intra-wound vancomycin in total hip and knee arthroplasty with evidence demonstrating lower infection rates with the use of vancomycin. However, all 6 studies included in the meta-analysis were retrospective case series with Level-III evidence. Xu H et al published their meta-analysis of 4607 patients reporting a reduced PJI rate with the use of vancomycin but a higher incidence of local wound complications and superficial surgical site infections (24). Our trial had similar findings of a higher incidence of minor complications such as persistent wound drainage and stitch abscesses in the study group.

Although previous studies attempted to evaluate the efficacy of intra-wound vancomycin, most were either underpowered due to the relatively low incidence of PJI, or suffered from low quality evidence secondary to retrospective study designs or lack of randomization. To the best of our knowledge, this study is the first prospective randomized control trial on the use of intra-wound vancomycin in primary TKA.

### Effect of intra-wound vancomycin on PJI/SSI

In this trial, the use of intra-wound vancomycin powder in primary TKA failed to show a reduction in the incidence of PJI/SSI. Previously published studies also showed no decrease in the incidence of PJI with local vancomycin use (29,30)(31). On the contrary, Winkler et al showed a statistically significant decrease in PJI in primary TKA. In the same study, however, there was only a trend towards decreased PJI with local antibiotic administration in THA and revision TKA/THA without statistical significance (32). The main limitation of this report was the retrospective study design and heterogeneity of the study population which included both primary and revision knee or hip arthroplasty, with varying intra-operative protocols.

Most studies reporting reduced infection rates with the use of intra-wound vancomycin were retrospective (33,34). Otte et al in their study of 1640 patients showed that 1g of intra-wound vancomycin significantly reduced the incidence of PJI in both primary and revision scenarios for both hips and knees (35). Patel et al also reported that intra-wound vancomycin was both safe and effective in reducing rates of early PJI in both primary hip and knee arthroplasties (36).

A meta-analysis by Peng et al, which included 4512 patients in 9 studies, also recommended the use of vancomycin powder to reduce the incidence of PJI without modifying the bacterial spectrum (37). A recent systematic review of 3371 patients did not demonstrate a significant reduction in PJI in patients receiving vancomycin (38). This was similar to the findings of the current trial.

This trial also showed a statistically significant increase in minor wound complications such as persistent wound drainage and stitch abscesses with the use of vancomycin. However, this should be interpreted with caution, as the trial may be underpowered to evaluate minor complications. Similar wound complications were also reported in previously published studies (20,24,26,33). The exact cause of this has not been determined but has been attributed to the crystalline nature of the vancomycin salt, low pH of the vancomycin solution or as a part of the body’s inflammatory response to the vancomycin which leads to seroma formation and subsequent wound complications (39). The aseptic wound complications in our study were managed using repeated sterile wound dressings without the routine use of antibiotics. Management of persistent wound drainage and breakdown can vary with some institutes opting for wound debridement and secondary closure in the operation theatre (20,33).

### Use of intra-wound vancomycin in diabetics and other high-risk groups

The use of vancomycin did not appear to significantly influence SSI/PJI rates in diabetics versus non-diabetics in the study population. There was no difference in outcomes based on the BMI classification of patients.

The strengths of this study include the prospective randomised study design, the trial performed at a single high-volume arthroplasty institute with a standardised protocol followed in all cases and a very low attrition during the follow-up period.

This study has some limitations. Firstly, this study is powered to detect a 1.7% difference in the PJI rates between the study and control groups. The trial is not adequately powered to detect smaller differences in PJI rates. The study may be underpowered for the minor complications such as prolonged wound drainage and delayed wound healing. This calls for large-volume multi-centre RCTs, with uniform protocols in peri-operative management. Registry-based studies will be high-powered but subject to bias due to varying institutional protocols in the asepsis procedures and antibiotic policies. Secondly, the results in this trial are based on data from a single institute. Although the surgical technique, aseptic precautions and antibiotic policy have been standardised across the study population, the findings may not be generalizable to other institutions dissimilar to ours. Other limitations include sub-analysis of other factors potentially contributing to PJI rate such as smoking or individual comorbidities, which were not independently evaluated.

## CONCLUSION

Intra-wound vancomycin powder application does not appear to reduce PJI/SSI rate in primary total knee arthroplasties, including in high-risk groups. The use of intra-wound vancomycin was associated with an increase in postoperative aseptic wound complications such as persistent wound drainage. Intra-wound vancomycin may not be effective in reducing the rate of PJI in primary TKA.

## Data Availability

All data produced in the present study are available upon reasonable request to the authors

## REFERENCES

1. Ackerman IN, Bohensky MA, Zomer E, Tacey M, Gorelik A, Brand CA, et al. The projected burden of primary total knee and hip replacement for osteoarthritis in Australia to the year 2030. BMC Musculoskelet Disord. 2019 Feb;20(1):90.

2. Kim TW, Kang S-B, Chang CB, Moon S-Y, Lee Y-K, Koo K-H. Current Trends and Projected Burden of Primary and Revision Total Knee Arthroplasty in Korea Between 2010 and 2030. J Arthroplasty. 2021;36(1):93–101.

3. Kurtz S, Ong K, Lau E, Mowat F, Halpern M. Projections of primary and revision hip and knee arthroplasty in the United States from 2005 to 2030. J Bone Joint Surg Am. 2007 Apr;89(4):780–5.

4. Springer BD, Cahue S, Etkin CD, Lewallen DG, McGrory BJ. Infection burden in total hip and knee arthroplasties: an international registry-based perspective. Arthroplast today. 2017 Jun;3(2):137–40.

5. Kapadia BH, Berg RA, Daley JA, Fritz J, Bhave A, Mont MA. Periprosthetic joint infection. Lancet (London, England). 2016 Jan;387(10016):386–94.

6. Kurtz SM, Lau E, Schmier J, Ong KL, Zhao K, Parvizi J. Infection burden for hip and knee arthroplasty in the United States. J Arthroplasty. 2008 Oct;23(7):984–91.

7. Clohisy JC, Calvert G, Tull F, McDonald D, Maloney WJ. Reasons for revision hip surgery: a retrospective review. Clin Orthop Relat Res. 2004 Dec;(429):188–92.

8. Fehring TK, Odum S, Griffin WL, Mason JB, Nadaud M. Early failures in total knee arthroplasty. Clin Orthop Relat Res. 2001 Nov;(392):315–8.

9. Vessely MB, Whaley AL, Harmsen WS, Schleck CD, Berry DJ. The Chitranjan Ranawat Award: Long-term survivorship and failure modes of 1000 cemented condylar total knee arthroplasties. Clin Orthop Relat Res. 2006 Nov;452:28–34.

10. Premkumar A, Kolin DA, Farley KX, Wilson JM, McLawhorn AS, Cross MB, et al. Projected Economic Burden of Periprosthetic Joint Infection of the Hip and Knee in the United States. J Arthroplasty. 2021 May;36(5):1484–1489.e3.

11. Kurtz SM, Lau E, Watson H, Schmier JK, Parvizi J. Economic burden of periprosthetic joint infection in the United States. J Arthroplasty. 2012 Sep;27(8 Suppl):61–5.e1.

12. Dodson V, Majmundar N, Swantic V, Assina R. The effect of prophylactic vancomycin powder on infections following spinal surgeries: a systematic review. Neurosurg Focus. 2019 Jan;46(1):E11.

13. Caroom C, Tullar JM, Benton EGJ, Jones JR, Chaput CD. Intrawound vancomycin powder reduces surgical site infections in posterior cervical fusion. Spine (Phila Pa 1976). 2013 Jun;38(14):1183–7.

14. Bakhsheshian J, Dahdaleh NS, Lam SK, Savage JW, Smith ZA. The use of vancomycin powder in modern spine surgery: systematic review and meta-analysis of the clinical evidence. World Neurosurg. 2015 May;83(5):816–23.

15. Yan H, He J, Chen S, Yu S, Fan C. Intrawound application of vancomycin reduces wound infection after open release of post-traumatic stiff elbows: a retrospective comparative study. J shoulder Elb Surg. 2014 May;23(5):686–92.

16. Wukich DK, Dikis JW, Monaco SJ, Strannigan K, Suder NC, Rosario BL. Topically Applied Vancomycin Powder Reduces the Rate of Surgical Site Infection in Diabetic Patients Undergoing Foot and Ankle Surgery. Foot ankle Int. 2015 Sep;36(9):1017–24.

17. Johnson JD, Nessler JM, Horazdovsky RD, Vang S, Thomas AJ, Marston SB. Serum and Wound Vancomycin Levels After Intrawound Administration in Primary Total Joint Arthroplasty. J Arthroplasty. 2017 Mar;32(3):924–8.

18. Burdon DW. Principles of antimicrobial prophylaxis. World J Surg. 1982 May;6(3):262–7.

19. Fleischman AN, Austin MS. Local Intra-wound Administration of Powdered Antibiotics in Orthopaedic Surgery. J bone Jt Infect. 2017;2(1):23–8.

20. Hanada M, Nishikino S, Hotta K, Furuhashi H, Hoshino H, Matsuyama Y. Intrawound vancomycin powder increases post-operative wound complications and does not decrease periprosthetic joint infection in primary total and unicompartmental knee arthroplasties. Knee Surg Sports Traumatol Arthrosc. 2019 Jul;27(7):2322–7.

21. Parvizi J, Gehrke T, Chen AF. Proceedings of the International Consensus on Periprosthetic Joint Infection. Bone Joint J. 2013 Nov;95-B(11):1450–2.

22. Parvizi J, Tan TL, Goswami K, Higuera C, Della Valle C, Chen AF, et al. The 2018 Definition of Periprosthetic Hip and Knee Infection: An Evidence-Based and Validated Criteria. J Arthroplasty. 2018 May;33(5):1309–1314.e2.

23. Ghanem E, Heppert V, Spangehl M, Abraham J, Azzam K, Barnes L, et al. Wound management. J Orthop Res Off Publ Orthop Res Soc. 2014 Jan;32 Suppl 1:S108–19.

24. Xu H, Yang J, Xie J, Huang Z, Huang Q, Cao G, et al. Efficacy and safety of intrawound vancomycin in primary hip and knee arthroplasty. Bone Joint Res. 2020 Nov;9(11):778–88.

25. Makris K, Spanou L. Acute Kidney Injury: Definition, Pathophysiology and Clinical Phenotypes. Clin Biochem Rev. 2016 May;37(2):85–98.

26. Kang DG, Holekamp TF, Wagner SC, Lehman RAJ. Intrasite vancomycin powder for the prevention of surgical site infection in spine surgery: a systematic literature review. Spine J. 2015 Apr;15(4):762–70.

27. Tubaki VR, Rajasekaran S, Shetty AP. Effects of using intravenous antibiotic only versus local intrawound vancomycin antibiotic powder application in addition to intravenous antibiotics on postoperative infection in spine surgery in 907 patients. Spine (Phila Pa 1976). 2013 Dec;38(25):2149–55.

28. Heckmann ND, Mayfield CK, Culvern CN, Oakes DA, Lieberman JR, Della Valle CJ. Systematic Review and Meta-Analysis of Intrawound Vancomycin in Total Hip and Total Knee Arthroplasty: A Call for a Prospective Randomized Trial. J Arthroplasty. 2019 Aug;34(8):1815–22.

29. Koutalos AA, Drakos A, Fyllos A, Doxariotis N, Varitimidis S, Malizos KN. Does Intra-Wound Vancomycin Powder Affect the Action of Intra-Articular Tranexamic Acid in Total Joint Replacement? Microorganisms. 2020 May;8(5).

30. Yavuz IA, Oken OF, Yildirim AO, Inci F, Ceyhan E, Gurhan U. No effect of vancomycin powder to prevent infection in primary total knee arthroplasty: a retrospective review of 976 cases. Knee Surg Sports Traumatol Arthrosc. 2020 Sep;28(9):3055–60.

31. Khatri K, Bansal D, Singla R, Sri S. Prophylactic intrawound application of vancomycin in total knee arthroplasty. J Arthrosc Jt Surg [Internet]. 2017;4(2):61–4. Available from: https://www.sciencedirect.com/science/article/pii/S2214963517300330

32. Winkler C, Dennison J, Wooldridge A, Larumbe E, Caroom C, Jenkins M, et al. Do local antibiotics reduce periprosthetic joint infections? A retrospective review of 744 cases. J Clin Orthop trauma. 2018 Mar;9(Suppl 1):S34–9.

33. Dial BL, Lampley AJ, Green CL, Hallows R. Intrawound Vancomycin Powder in Primary Total Hip Arthroplasty Increases Rate of Sterile Wound Complications. Hip pelvis. 2018 Mar;30(1):37–44.

34. Xu X, Zhang X, Zhang Y, Chen C, Yu H, Xue E. Role of intra-wound powdered vancomycin in primary total knee arthroplasty. Orthop Traumatol Surg Res. 2020 May;106(3):417–20.

35. Otte JE, Politi JR, Chambers B, Smith CA. Intrawound Vancomycin Powder Reduces Early Prosthetic Joint Infections in Revision Hip and Knee Arthroplasty. Surg Technol Int. 2017 Feb;30:284–9.

36. Patel NN, Guild GN 3rd, Kumar AR. Intrawound vancomycin in primary hip and knee arthroplasty: a safe and cost-effective means to decrease early periprosthetic joint infection. Arthroplast today. 2018 Dec;4(4):479–83.

37. Peng Z, Lin X, Kuang X, Teng Z, Lu S. The application of topical vancomycin powder for the prevention of surgical site infections in primary total hip and knee arthroplasty: A meta-analysis. Orthop Traumatol Surg Res. 2021 Jun;107(4):102741.

38. Wong MT, Sridharan SS, Davison EM, Ng R, Desy NM. Can Topical Vancomycin Prevent Periprosthetic Joint Infection in Hip and Knee Arthroplasty? A Systematic Review. Clin Orthop Relat Res. 2021 Aug;479(8):1655–64.

39. Hoelen DWM, Tjan DHT, van Vugt R, van der Meer YG, van Zanten ARH. Severe local vancomycin induced skin necrosis. Vol. 64, British journal of clinical pharmacology. England; 2007. p. 553–4.

